# Long term impact of coal mine fire smoke on lung mechanics in exposed adults

**DOI:** 10.1101/2020.10.14.20213009

**Authors:** Nicolette R Holt, Caroline X. Gao, Brigitte M Borg, David Brown, Jonathan C Broder, Jillian Ikin, Annie Makar, Thomas McCrabb, Kris Nilsen, Bruce R Thompson, Michael J. Abramson

**Author notes:** **Correspondence to:** Professor Michael Abramson, School of Public Health & Preventive Medicine, Monash University, 553 St Kilda Rd, Melbourne, Vic 3004, Australia.

## Abstract

In 2014, a six-week long fire at the Hazelwood open cut coal mine exposed residents in the adjacent town of Morwell to high concentrations of fine particulate matter with an aerodynamic diameter <2.5μm (PM_2.5_). The long-term health consequences are being evaluated as part of the Hazelwood Health Study (HHS).

Approximately 3.5 to 4 years after the mine fire, adults from Morwell (n=346) and the comparison town Sale (n=173) participated in the longitudinal Respiratory Stream of the HHS. Individual fire-related PM_2.5_ exposure was retrospectively modelled. Lung mechanics were assessed using the forced oscillation technique (FOT), which utilises pressure waves to measure respiratory system resistance (Rrs) and reactance (Xrs). Multivariate linear regression was used to evaluate associations between PM_2.5_ and transformed Rrs5, area under the reactance curve (AX5) and Xrs5 controlling for key confounding factors.

There were clear dose-response relationships between increasing mine fire PM_2.5_ and worsening lung mechanics, including a reduction in post-bronchodilator Xrs5 and an increase in AX5. A 10 μg/m^3^ increase in mine fire related PM_2.5_ was associated with a 0.015 (95%CI: 0.004, 0.027) reduction in exponential(Xrs5) post bronchodilator, which was comparable to 4.7 years of aging. Similarly, the effect of exposure was associated with a 0.072 (0.005, 0.138) increase in natural log(AX5) post-bronchodilator, equivalent to 3.9 years of aging.

This is the first study using FOT in adults evaluating long term respiratory outcomes after a medium-term ambient PM_2.5_ exposure to coal mine fire smoke. These results should inform public health policies and planning for future events.

## INTRODUCTION

Ambient air particulate matter (PM) exposure, from sources including vehicle exhaust, industry, biomass fuels and wildfires, collectively account for an estimated 7.5% of all deaths globally in 2016.[1] In particular, fine PM with an aerodynamic diameter < 2.5μm (PM_2.5_) infiltrates deep into the peripheral lung. Short-term (days) exposure to PM_2.5_ has been shown to be associated with cardiovascular and respiratory morbidity and mortality.[2,3] Guo et al[4] showed in a large cohort study that long-term ambient PM_2.5_ exposure was consistently associated with reduced lung function, accelerated annual lung function decline and an increased risk of developing chronic obstructive pulmonary disease (COPD) in adults. Similarly in a review, Li et al[5] showed an association with long-term exposure to ambient air pollution levels and increased incidence of respiratory symptoms in children.

The long-term sequelae of fine particle exposures on lung function, particularly from medium exposure episodes (weeks to months) such as landscape fires, have not been well characterised. Studies of wildfires predominantly use secondary data such as hospitalization and emergency presentations to identify respiratory associations.[6] Though long-term exposure to indoor coal burning has been found to be associated with worsening respiratory symptoms, reduced lung function and chronic obstructive pulmonary disease in adults - much remains unknown regarding the impact of coal mine fires on human lung health.[6] Addressing the gaps in the current available evidence is critical given the increasing incidence of catastrophic wildfires globally attributable to climate change.[7]

The forced oscillation technique (FOT) is a methodology used to measure lung mechanics. FOT may be able to detect early changes in peripheral airway function that spirometry cannot.[8] To our knowledge, no study has assessed the long-term impact of PM_2.5_ from exposure to coal mine fires, wildfires, or biomass fuel smoke in adults using FOT.

In February 2014, embers from nearby bush fires started a fire in the Hazelwood open-cut brown coal mine, located in the Latrobe Valley, south-eastern Australia. It was an unprecedented event that generated significant air pollution from coal mine fire smoke over six weeks, particularly affecting residents in the adjacent town of Morwell. The most exposed population at the time numbered approximately 14,000.[9] This resulted in considerable community concerns about the potential long-term health effects of smoke exposure.

The Hazelwood Health Study (www.hazelwoodhealthstudy.org.au) was established to investigate potential health effects in people who were exposed to smoke from the mine fire. The Hazelinks stream of the Hazelwood Health Study utilised hospital emergency presentations and admissions data to show that hospitalisation for respiratory conditions increased during the first month of the mine fire.[10] The Adult Survey stream of the Hazelwood Health Study[11] compared self-reported health outcomes between the most exposed community and an unexposed sample more than two years after the event. The Survey found increasing risks of respiratory symptoms, particularly cough, phlegm and wheeze, related to the mine fire exposure.[12] This analysis aimed to further investigate the association between exposure to mine fire smoke and long-term lung function as assessed by FOT, 3.5 to four years after the event.

## METHODS

### Study design and setting

The Respiratory Stream of the Hazelwood Health Study is a longitudinal follow-up study of selected participants from the Adult Survey.[11,13]. The study was conducted between August and December 2017 in Morwell (exposed), and between January and March 2018 in the nearby town of Sale (unexposed). Study data were collected and managed using REDCap (Research Electronic Data Capture)[14] electronic data capture tools hosted at Monash University, Australia.

### Participant eligibility and recruitment

Participants were eligible for the Respiratory Stream of the Hazelwood Health Study if they had completed the Adult Survey, were at least 18 years of age on 9 February 2014 and had lived in the study area at the time of the mine fire. Adult Survey participants were excluded from the Respiratory Stream if they had specified no further contact, were of unknown age or sex, or were aged over 90 years. Participants were further excluded where a contraindication to spirometry was identified – including recent surgery, myocardial infarction, pneumothorax, pulmonary embolism, open pulmonary tuberculosis or known aneurysms.[15] A target sample size of 339 from Morwell and 170 from Sale was derived based on the ability to detect a 5ml/year or greater FEV_1_ decline in exposed compared with non-exposed participants using a two-sample t-test with a two-sided p-value of 0.05 and 80% power. A weighted random sample (to correct for lower response rate in some subgroups of participants, such as young people) of 1,346 Adult Survey participants was invited for assessment of their respiratory function. Participants reporting an asthma attack or current asthma medication use in the Adult Survey were oversampled (40%) to provide ability for further evaluation in an asthmatic sample. Invitation to participate was by mail, email and/or SMS, and recruitment continued until the target sample size was achieved (see Figure 1).

### Participant characteristics

Participant characteristics such as age, sex, ethnicity, employment status and smoking history were collected via questionnaires. Participants were classified as non-smokers (<100 cigarettes in their lifetimes), ex-smokers (current non-smokers with >100 cigarettes in their lifetimes) or current smokers.[16] Height and weight were measured by trained personnel during the study visit. Education level and occupational exposures (employment in dusty or polluted environments for at least six months) were obtained from the Adult Survey.[11] Self-reported asthma status was captured via a modified European Community Respiratory Health Survey questionnaire.[17] Participants were identified as having spirometry consistent with COPD if post bronchodilator (BD) FEV_1_/FVC < lower limit of normal (5^th^ percentile) using Global Lung Initiative spirometry reference values.[18]

### Exposure assessment

Retrospective modelling of the spatial and temporal distribution of mine fire-related PM_2.5_ concentrations by the Australian Commonwealth Scientific and Industrial Research Organisation (CSIRO) Oceans & Atmosphere[19,20] was used due to the absence of ground-level air pollution monitoring at the beginning of the mine fire. Individual level mean daily PM_2.5_ exposures over the mine fire period (51 days, between 9 February to 31 March, 2014) were estimated through linking time-location diary data (reported in the Adult Survey) with the modelled fire-related PM_2.5_ exposure data as described by Johnson et al.[12]

### Clinical outcome measures

Respiratory testing was performed by the same trained respiratory scientists at both sites using standard operating procedures in line with current respiratory measurement standards where available. Spirometry was measured using the EasyOne Pro Lab Respiratory Analysis System (ndd Medical Technologies AG, Zürich, Switzerland) in line with international standards.[21] Forced Oscillation Technique (FOT) parameters were measured using the Tremoflo C-100device (Thorasys, Montreal, Canada) in line with standards current at time of testing.[22] Parameters reported for FOT included respiratory system resistance and reactance at a frequency of 5Hz (Rrs5 and Xrs5 respectively), the area under the reactance curve (AX5) and resonant frequency (Fres). Data were imputed[23] where acceptability criteria were not met or coherence <0.80 for 5Hz or <0.90 at 11 or 19Hz.[22] Tests were performed before and 10 minutes after administration of a short acting bronchodilator (300μg salbutamol). Bronchodilator use in the previous 24 hours was recorded, as bronchodilators were unable to be withheld prior to assessment due to ethical reasons.

### Statistical methods

Statistical weighting was developed and applied to all analyses to correct for over-sampling of asthmatics as well as possible attrition bias from the Adult Survey to clinical follow-up, see details in the online supplement. Descriptive statistics were used to compare patient characteristics and clinical outcomes between non-exposed Sale participants as well as the tertiles of PM_2.5_ exposure level in Morwell (low, medium or high exposure). Crude statistical significance was assessed using Pearson chi-squared tests for categorical measures and t-tests for continuous measures.

Multivariate linear regression models were fitted to analyse the association between mean PM_2.5_ exposure and outcomes, controlling for key confounders including age, height, weight, sex, smoking status, self-reported asthma and/or COPD, employment, education level and occupational exposure. Standardised z-scores and %predicted[24] for FOT outcome variables were not used in the analysis due to poor regression model fit and high proportions of participants outside of reference prediction range (mostly due to older age and heavier weight). Therefore, possible outcome transformation methods and nonlinear associations were explored using both Box-Cox transformation and fractional polynomial regression models. The best outcome transformation methods were identified as logarithmic transformations for Rrs5, AX5 and Fres and exponential transformation for Xrs5. Additional non-linearity was not observed between transformed outcomes and predicators such as age, weight and height. Missing data were addressed using multiple imputation using chained equations. Due to the lack of a low or no exposure sample in Morwell, as well as possible differences between Morwell and Sale participants, two sets of regression models were carried out for each outcome variable: one model including a binary variable indicating township of participant (Morwell or Sale), and the other model excluding this variable. Sensitivity analyses were performed with unweighted and complete case models. Statistical analyses were performed using Stata version 15 (Stata Corporation, College Station, Texas 2015).

### Ethical considerations

The Monash University Human Research Ethics Committee (MUHREC) approved the Hazelwood Health Study: Cardiovascular and Respiratory Streams (approval number 1078). All participants provided written informed consent.

## RESULTS

### Participant characteristics and PM_2.5_ exposure

This cross-sectional analysis included all participants in the first round of Respiratory Stream data collection, which comprised a total of 519 participants (346 from Morwell, and 173 from Sale). Refer to **Figure 1** for flow of participants.

**Table 1** shows the participant characteristics by exposure level to mine fire smoke. The mean (standard deviation; SD) PM_2.5_ exposure levels for non-exposed (Sale) and for Morwell (low, medium and high exposure groups) were 0.1 (0.4), 5.9 (1.8), 11.5 (1.5) and 27.8 (10.3) μg/m^3^, respectively. There were differences between exposure groups for gender distribution and weight, with those in the high exposure group having a higher proportion of males and heavier weight. Other participant characteristics were comparable between exposure groups.

**Table 1:**
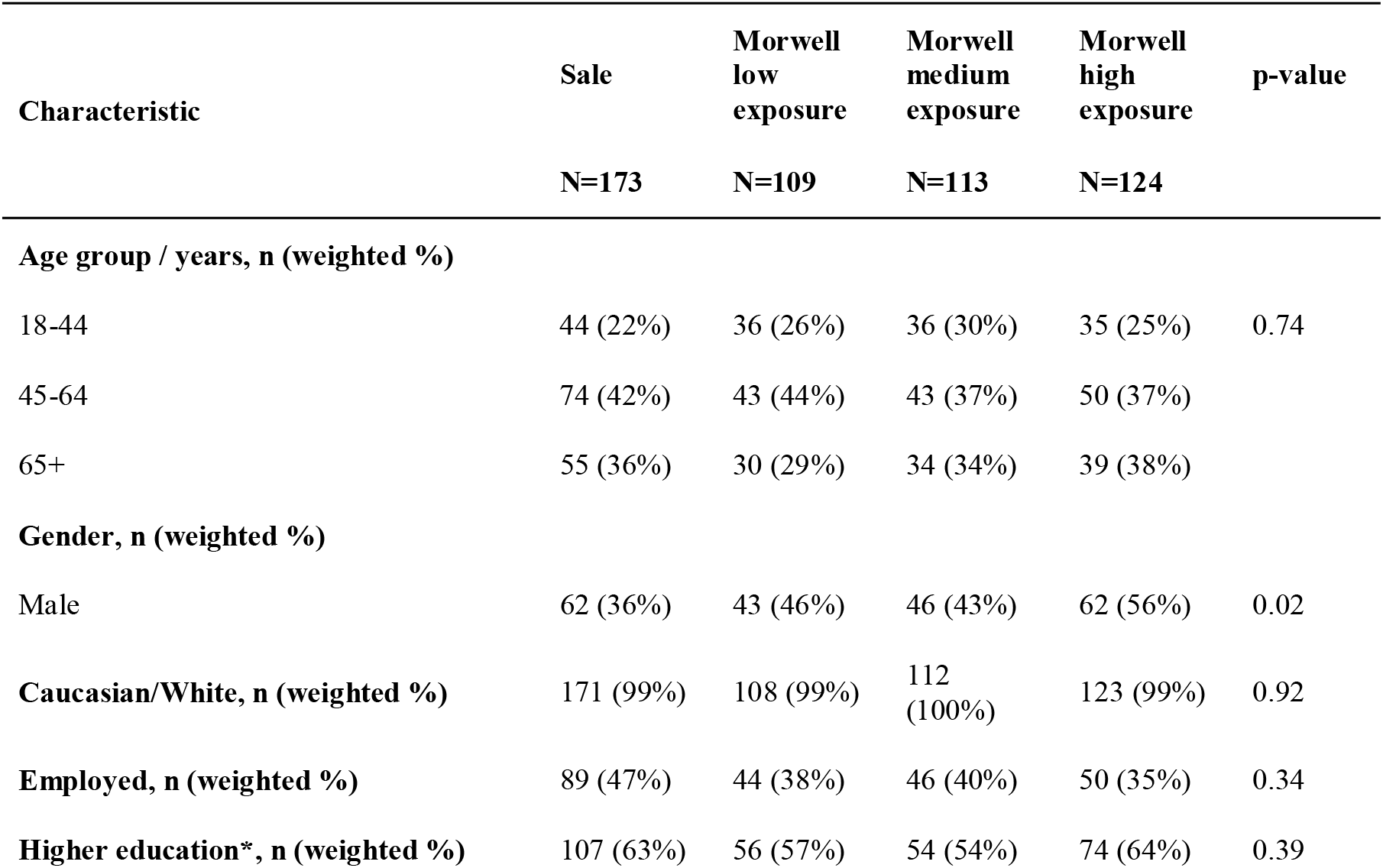
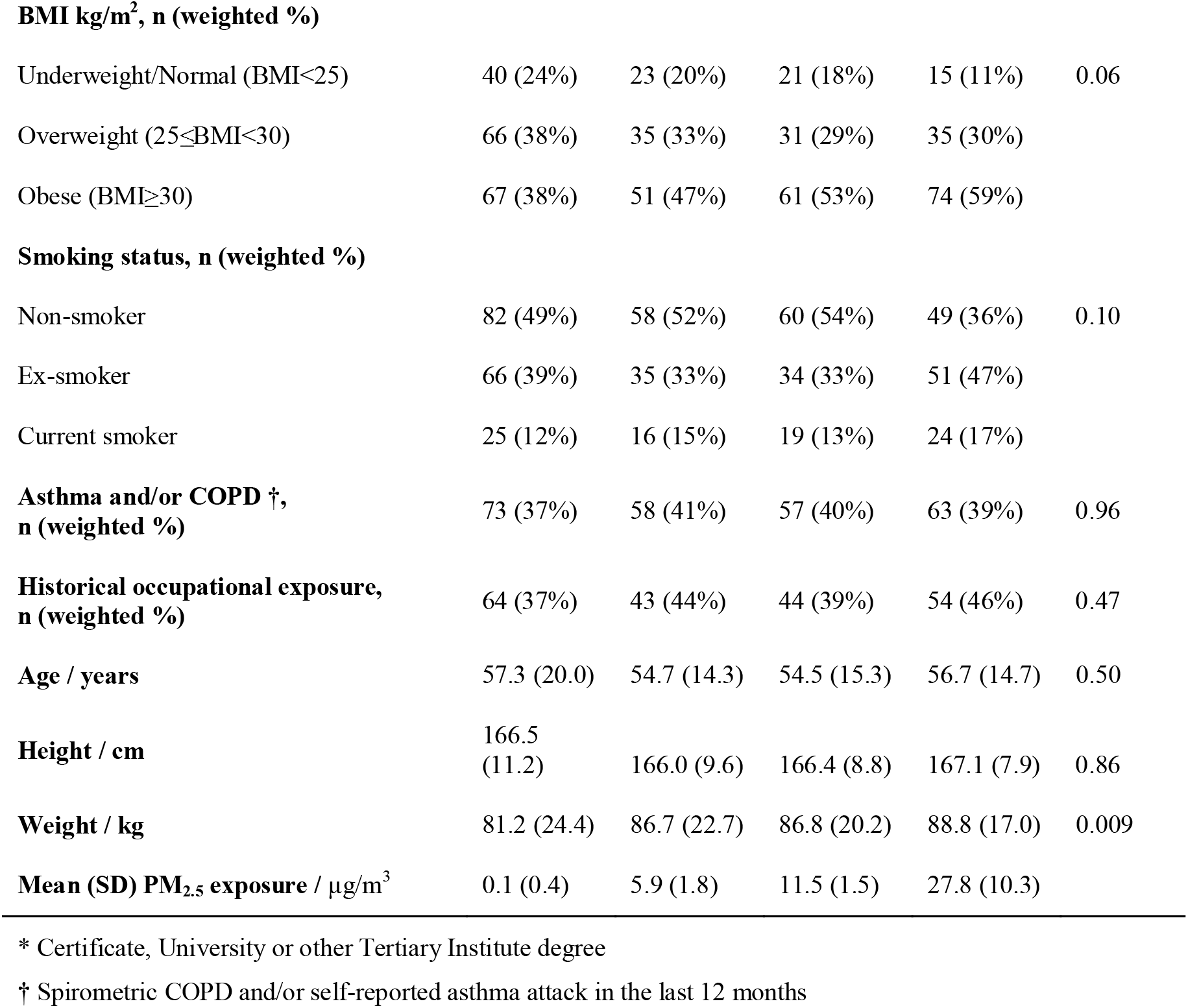
Participant characteristics by exposure group.

### PM_2.5_ exposure and lung function

Forced Oscillation Technique variables are dependent on sex, age, height and weight – hence unadjusted results lack meaning and were not included in the analysis. As shown in **Figures 2A and 2B** and **Table S1**, all outcome variables were skewed and displayed slightly larger variation in baseline compared to post bronchodilator outcomes. A clear dose response pattern was observed between exposure level and FOT outcomes. Results from multivariate linear regression analysis (**Table 2**) revealed a negative association between increasing mine fire related PM_2.5_ exposure and post bronchodilator reactance at 5Hz, with Morwell included or excluded as a predictor. With Morwell excluded as a predictor, a 10 μg/m^3^ increase in mine fire related PM_2.5_ was associated with 0.015 reduction in post bronchodilator exponential transformed Xrs5. This was equivalent to 4.7 years of aging estimated in the regression model (see Table S2). When Morwell was excluded as a predictor, regression analysis suggested that increased exposure to mine fire related PM_2.5_ was associated with increased area under the post bronchodilator reactance curve (AX5). The effect of exposure was associated with a 0.072 increase in ln(AX5) post-bronchodilator; being equivalent to 3.9 years of aging (see Table S3). More detailed regression results for post bronchodilator Xrs5 and AX5 are shown in supplementary **Tables S2 to S3**. Sensitivity analyses (results not shown) suggest that un-weighted and complete case results were consistent with main findings.

**Table 2.** Summary table for multivariate linear regressions of FOT parameters – regression coefficients (β) and 95% confidence intervals.

| | Mean exposure model (10 $\mu\text{g}/\text{m}^3$)<br>Including Morwell as predictor | | Mean exposure model (10 $\mu\text{g}/\text{m}^3$)<br>Excluding Morwell as predictor | |
| --- | --- | --- | --- | --- |
| | β-Coef (95% CI) | p-value | β-Coef (95% CI) | p-value |
| **Baseline\*** |  |  |  |  |
| Baseline ln(Rrs5) | -0.003 (-0.035, 0.029) | 0.87 | -0.001 (-0.028, 0.026) | 0.95 |
| Baseline exp(Xrs5) | -0.009 (-0.024, 0.006) | 0.23 | -0.008 (-0.020, 0.005) | 0.23 |
| Baseline ln(AX5) | 0.030 (-0.056, 0.116) | 0.50 | 0.038 (-0.034, 0.109) | 0.31 |
| Baseline ln(Fres) | 0.001 (-0.030, 0.032) | 0.97 | 0.006 (-0.019, 0.032) | 0.62 |
| **Post BD†** |  |  |  |  |
| Post BD ln(Rrs5) | 0.011 (-0.018, 0.041) | 0.45 | 0.012 (-0.013, 0.036) | 0.34 |
| Post BD exp(Xrs5) | **-0.018 (-0.032, -0.003)** | **0.015** | **-0.015 (-0.027, -0.004)** | **0.011** |
| Post BD ln(AX5) | 0.063 (-0.017, 0.144) | 0.12 | **0.072 (0.005, 0.138)** | **0.034** |
| post BD ln(Fres) | 0.017 (-0.010, 0.045) | 0.22 | 0.021 (-0.001, 0.044) | 0.07 |

\* Regression models adjusted for age, gender, height, weight, employment, education, smoking status, asthma and COPD status and work exposure and whether participants had bronchodilator prior to the test. Missing data, including 44 records for baseline Rrs5, baseline Xrs5 and baseline AX5; 48 records for baseline Fres; 6 records for education level, were imputed using multiple imputation with chained equations.
† Regression models adjusted for age, gender, height, weight, employment, education, smoking status, asthma and COPD status and work exposure. Missing data, including 41 records for post BD Rrs5, post BD Xrs5 and post BD AX5; 42 records for post BD Fres; 6 records for education level, were imputed using multiple imputation with chained equations.
Note: exp- exponential; ln- natural log

## DISCUSSION

Assessment of participants nearly four years after the Hazelwood coal mine fire revealed an association between medium term mine fire related PM_2.5_ exposure and more negative respiratory system reactance (Xrs5), specifically measured after administration of bronchodilator. To the best of our knowledge, this represents the first study using FOT analysis in adults to evaluate longer term respiratory function after a medium term PM_2.5_ exposure related to coal mine fire smoke.

The mechanism for the more negative reactance (a marker of the compliance of the respiratory system) is unclear. Previous studies of long-term exposure to air pollution and PM_2.5_ have shown associations with increased respiratory morbidity and airflow obstruction.[4,6,25-27] Separately, it has been shown that measurements of reactance at 5-6Hz via FOT are sensitive to airway closure[28-31] and expiratory flow limitation[32-34] in subjects with obstruction. A possible mechanism for the association seen between medium term exposure PM_2.5_ and Xrs5 in this study may be early peripheral airway changes that occur with airflow limitation or accelerated lung aging.

Interestingly, the association between PM_2.5_ and Xrs5 was only observed in the post-bronchodilator data. A possible explanation for this finding is that participants were recruited from a general population with varying states of lung health and by assessing participants post-bronchodilator, variability of bronchomotor tone was minimised across participants[35-37] allowing assessment of fixed pulmonary abnormalities. That is, the assessment of the relationship between PM_2.5_ and Xrs5 could be undertaken without the confounding effects of bronchomotor tone.

Importantly, these findings in adults are similar to the findings in children within the Hazelwood Health Study Early Life Follow up (ELF) stream. Shao and colleagues[38] demonstrated that infant or *in utero* exposures to coal mine fire emissions were associated with long-term impairment of lung reactance, with increased average PM_2.5_ being significantly associated with worsening area under the reactance curve - a complementary parameter in the evaluation of reactance.[24]

The study has several strengths. Unlike observational studies that have used only secondary data (such as hospitalization) to assess respiratory health, this research has built upon previously collected hospitalisation[10] and self-reported symptom data[12] with the inclusion of objective measures of lung mechanics. A further strength of this study was the inclusion of individual estimates of PM_2.5_ exposure utilising a combination of detailed time-location diaries and spatially and temporally resolved modelling of PM_2.5_ concentrations based upon coal combustion and weather conditions.

However, the study also has some limitations. The study endeavoured to account for all relevant potential confounding factors in our analysis, such as age, gender, weight, BMI, education status, tobacco and occupational exposures. However, it is feasible that some of the observed results occurred by chance or were influenced by unknown confounding factors. Furthermore, at this stage in the study we only have cross-sectional data on lung mechanics. Future followup of the Hazelwood Health Study Respiratory Stream participants will better inform an investigation of the long-term implications of medium-duration coal mine fire-related smoke exposure on respiratory mechanics and lung health.

In conclusion, a clear dose response association was observed between medium-duration PM_2.5_ exposure levels from ambient coal mine fire smoke and a more negative respiratory system reactance in this cohort. This study adds new findings to the literature on the lung health effects of medium term PM_2.5_ exposure. These inform public health policy and planning for future coal mine fires or similar medium duration PM_2.5_ generating pollution events such as the recent megafires in Australia and the United States. Longitudinal data are required to confirm the findings of this study and to better understand the association of coal mine fire smoke and altered respiratory system reactance and potential accelerated lung aging in exposed populations.

## Data Availability

Intellectual property rights to the data are held by the Victorian Department of Health and Human Services. Parties interested in collaborating with the Hazelwood Health Study to use this data should contact the Principal Investigator

## Acknowledgments

We wish to thank Susan Denny, Kylie Sawyer, Shantelle Allgood and Kristina Thomas from the Monash University School of Rural Health, who assisted with the study. Most of all, the study team would like to acknowledge the contribution of all community members who have participated in the study to date.

## Financial support

The Hazelwood Health Study is funded by the Victorian Department of Health & Human Services (Australia). However, this paper presents the views of the authors and does not represent the views of the Department.

---

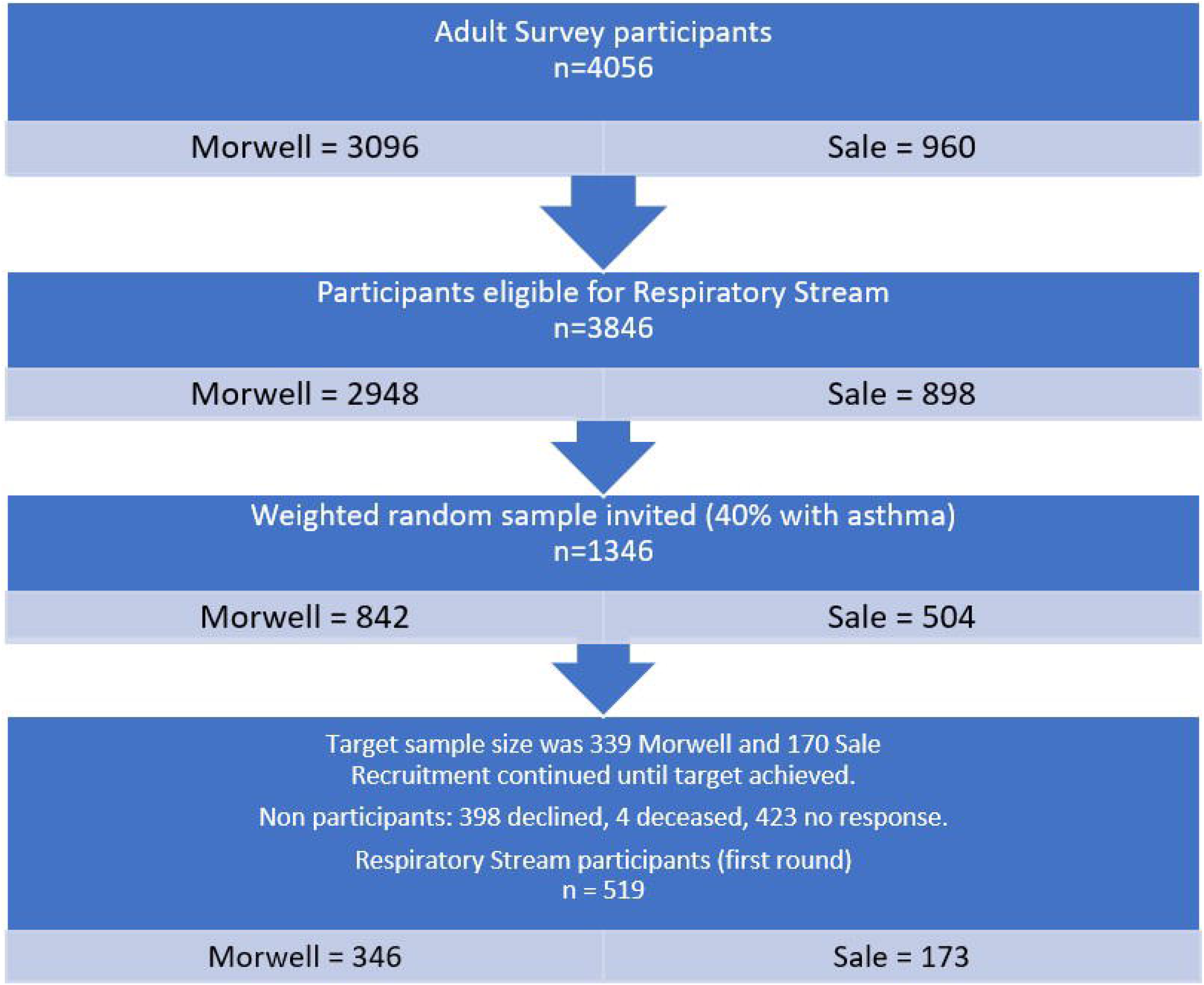

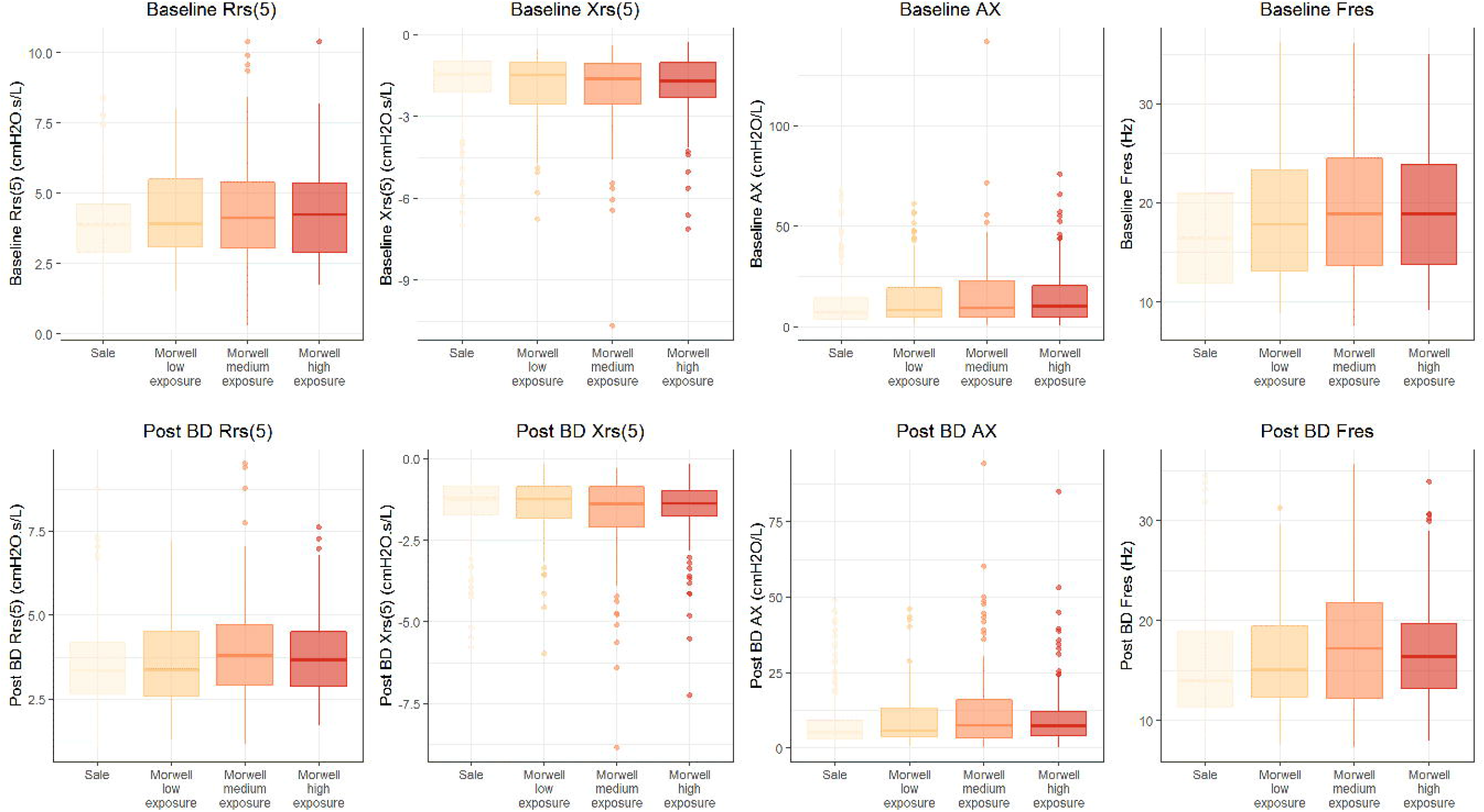

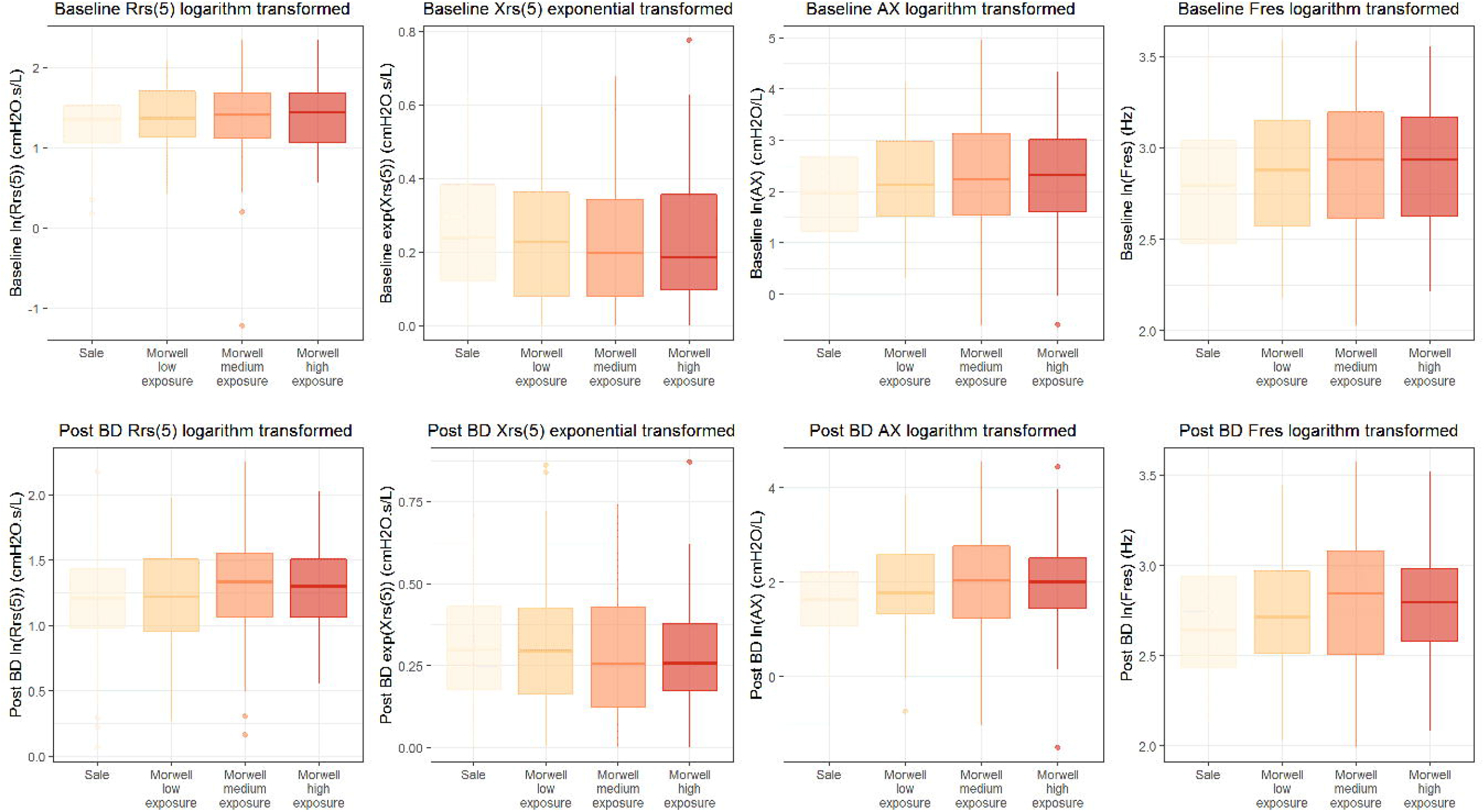

## Notes

### Competing Interest Statement

MA reports investigator initiated grants for unrelated research from Pfizer and Boehringer-Ingelheim. He has also undertaken an unrelated consultancy for Sanofi and received a speaker's fee from GSK. Other authors declare no conflicts of interest.

### Funding Statement

This work was funded by the Victorian Department of Health and Human Services (Victoria, Australia). The paper presents the views of the authors and does not represent the views of the Department.

### Author Declarations

The Monash University Human Research Ethics Committee (MUHREC) approved the Hazelwood Health Study: Cardiovascular and Respiratory Streams (approval number 1078).

