## supplement for "Long term impact of coal mine fire smoke on lung mechanics in exposed adults"

**Online Supplement Material**

***Statistical weighting***

Statistical weighting was developed to correct for oversampling of the asthmatic population as well as potential attrition bias. The over-sampling weight was determined simply by the inverse sampling probabilities of selection in each of the location and asthmatic status group [39]. The attrition weight was developed via inverse the modelled probability of attrition from a set of “luxury” variables collected in the Adult Survey that were potentially related to both attrition and outcome. The chosen variables included age, gender, education, employment, marital, smoking and drinking status as well as self-reported asthma, COPD and chronic cough. The final study weight was calculated as the product of the over-sampling and attrition weights.

**Table S1:** FOT parameters by exposure group.

| **Outcome variables** | **Sale** | **Morwell low exposure** | **Morwell medium exposure** | **Morwell high exposure** |  |
| --- | --- | --- | --- | --- | --- |
|  | **N=173** | **N=109** | **N=113** | **N=124** |  |
|  | **Weighted mean(SD)** | **Weighted mean(SD)** | **Weighted mean(SD)** | **Weighted mean(SD)** | **p-value** |
| **Baseline** |  |  |  |  |  |
| Baseline Rrs5 | 3.8 (1.8) | 4.0 (1.4) | 4.3 (1.6) | 4.1 (1.4) | 0.21 |
| Baseline ln(Rrs5) | 1.3 (0.5) | 1.3 (0.3) | 1.4 (0.4) | 1.3 (0.3) | 0.41 |
| Baseline Xrs5 | -1.6 (1.3) | -1.7 (1.0) | -1.8 (1.2) | -1.8 (1.1) | 0.40 |
| Baseline exp(Xrs5) | 0.3 (0.2) | 0.3 (0.1) | 0.2 (0.2) | 0.2 (0.1) | 0.51 |
| Baseline AX5 | 10.4 (14.1) | 12.5 (12.3) | 13.9 (14.5) | 14.1 (12.8) | 0.07 |
| Baseline ln(AX5) | 1.9 (1.2) | 2.1 (0.9) | 2.1 (1.0) | 2.2 (0.9) | 0.11 |
| Baseline Fres | 16.9 (7.6) | 18.3 (6.1) | 18.5 (6.0) | 19.1 (6.0) | 0.041 |
| Baseline ln(Fres) | 2.8 (0.4) | 2.8 (0.3) | 2.9 (0.3) | 2.9 (0.3) | 0.050 |
| **Post BD** |  |  |  |  |  |
| Post BD Rrs5 | 3.5 (1.7) | 3.5 (1.2) | 3.8 (1.4) | 3.7 (1.1) | 0.19 |
| Post BD ln(Rrs5) | 1.2 (0.5) | 1.2 (0.3) | 1.3 (0.4) | 1.2 (0.3) | 0.18 |
| Post BD Xrs5 | -1.4 (1.1) | -1.3 (0.8) | -1.5 (1.0) | -1.5 (0.8) | 0.39 |
| Post BD exp(Xrs5) | 0.3 (0.2) | 0.3 (0.2) | 0.3 (0.2) | 0.3 (0.1) | 0.21 |
| Post BD AX5 | 8.1 (11.7) | 8.8 (8.4) | 10.9 (11.5) | 9.8 (9.5) | 0.19 |
| Post BD ln(AX5) | 1.6 (1.2) | 1.8 (0.9) | 1.9 (0.9) | 1.9 (0.8) | 0.07 |
| post BD Fres | 15.4 (6.8) | 16.0 (4.9) | 17.1 (5.7) | 16.7 (4.3) | 0.08 |
| Post BD ln(Fres) | 2.7 (0.4) | 2.7 (0.3) | 2.8 (0.3) | 2.8 (0.2) | 0.047 |

**Table S2:** Regression models for post BD reactance - exp(Xrs5)

|  | **Including Morwell as predictor** | | **Excluding Morwell as predictor** | |
| --- | --- | --- | --- | --- |
| **Predictors** | **β-Coef (95% CI)** | **p-value** | **β-Coef (95% CI)** | **p-value** |
| **Exposure to PM_2.5_ (10 µg/m³)** | -0.018 (-0.032,-0.003) | 0.015 | -0.015 (-0.027,-0.004) | 0.011 |
| **Age (10 years)** | -0.032 (-0.041,-0.022) | <0.001 | -0.032 (-0.042,-0.023) | <0.001 |
| **Height** | 0.007 (0.005,0.010) | <0.001 | 0.007 (0.005,0.010) | <0.001 |
| **Weight** | -0.002 (-0.003,-0.001) | <0.001 | -0.002 (-0.003,-0.001) | <0.001 |
| **Male** | 0.027 (-0.017,0.072) | 0.23 | 0.029 (-0.015,0.073) | 0.20 |
| **Smoking status** |  |  |  |  |
| Non-smoker | Ref |  | Ref |  |
| Ex-smoker | -0.002 (-0.033,0.029) | 0.90 | -0.003 (-0.034,0.028) | 0.84 |
| Current smoker | -0.033 (-0.075,0.009) | 0.13 | -0.034 (-0.075,0.008) | 0.12 |
| **Morwell** | 0.015 (-0.021,0.051) | 0.41 | - | - |
| **Asthma and/or COPD †** | -0.059 (-0.089,-0.029) | <0.001 | -0.058 (-0.088,-0.029) | <0.001 |
| **Employed** | -0.014 (-0.046,0.018) | 0.40 | -0.015 (-0.046,0.017) | 0.38 |
| **Higher education*** | -0.005 (-0.034,0.023) | 0.72 | -0.006 (-0.034,0.023) | 0.68 |
| **Work exposure** | 0.003 (-0.030,0.036) | 0.87 | 0.003 (-0.030,0.036) | 0.84 |

* Certificate, University or other Tertiary Institute degree

† Spirometry consistent with COPD and/or self-reported asthma attack in the last 12 months

**Table S3:** Regression results for post BD area under the reactance curve - ln(AX5)

|  | **Including Morwell as predictor** | | **Excluding Morwell as predictor** | |
| --- | --- | --- | --- | --- |
| **Predictors** | **β-Coef (95% CI)** | **p-value** | **β-Coef (95% CI)** | **p-value** |
| **Exposure to PM_2.5_ (10 µg/m³)** | 0.063 (-0.017,0.144) | 0.12 | 0.072 (0.005,0.138) | 0.034 |
| **Age (10 years)** | 0.188 (0.136,0.239) | <0.001 | 0.185 (0.134,0.237) | <0.001 |
| **Height** | -0.042 (-0.054,-0.029) | <0.001 | -0.042 (-0.054,-0.030) | <0.001 |
| **Weight** | 0.015 (0.011,0.019) | <0.001 | 0.015 (0.011,0.019) | <0.001 |
| **Male** | 0.040 (-0.189,0.269) | 0.73 | 0.046 (-0.181,0.273) | 0.69 |
| **Smoking status** |  |  |  |  |
| Non-smoker | Ref |  | Ref |  |
| Ex-smoker | -0.005 (-0.174,0.164) | 0.96 | -0.009 (-0.177,0.159) | 0.92 |
| Current smoker | 0.164 (-0.076,0.404) | 0.18 | 0.161 (-0.079,0.400) | 0.19 |
| **Morwell** | 0.052 (-0.146,0.250) | 0.60 | - | - |
| **Asthma and/or COPD†** | 0.372 (0.207,0.537) | <0.001 | 0.373 (0.209,0.537) | <0.001 |
| **Employed** | 0.064 (-0.116,0.243) | 0.49 | 0.061 (-0.118,0.240) | 0.50 |
| **Higher education*** | 0.000 (-0.156,0.156) | 0.99 | -0.003 (-0.158,0.153) | 0.98 |
| **Work exposure** | -0.014 (-0.191,0.164) | 0.88 | -0.011 (-0.190,0.167) | 0.90 |

* Certificate, University or other Tertiary Institute degree

† Spirometry consistent with COPD and/or self-reported asthma attack in the last 12 months
